# Pregnancy outcomes from a restorative infertility treatment model: a single centre case series

**DOI:** 10.1101/2021.04.14.21251044

**Authors:** Gabriel James, Lucas A. Mclindon, Joshua Hatch, Ben W. Mol, Joseph V. Turner

## Abstract

**Background:** Infertility is a significant problem with multiple causes and a corresponding array of therapeutic options. In an era of increasing assisted reproductive treatments, few studies examine the role of conventional non-assisted reproductive treatments to address underlying behavioural, lifestyle and medical issues.

**Aim:** To assess outcomes from a conventional or non-assisted reproductive treatment approach

**Materials and Methods:** Retrospective case series of 162 couples that attended an Australian, hospital-based, multidisciplinary fertility clinic between 2005 and 2010.

**Results:** There were 58 live births for all couples giving a crude live birth rate of 35.4% over a 24-month analysis period. When adjusted by Kaplan-Meier method, a 57.4% cumulative live birth rate (CLBR) was achieved. Couples had a median 33.9 months duration of infertility and the median female age was 33.7. For the 74 couples with an unexplained infertility diagnosis, 32 achieved a live birth at a crude rate of 43.2% or 71.2% CLBR when adjusted by Kaplan-Meier method.

**Conclusions:** This observational data indicates that reproductive medicine should have a personalized approach in which alternatives for immediate IVF are considered.

## Introduction

Infertility, defined by the World Health Organisation (WHO) as the inability to conceive after 12 months of uncontracepted intercourse, affects 1 in 6 couples^1^. In Australia, the largest longitudinal study on women’s health showed that only 50-70% of subfertile couples seek advice and fewer access assisted reproductive therapies (ART)^2,3^. When couples do access care, there are further challenges outlining an accurate prognosis^4^.

Professional bodies recommend specific counselling about chances of natural conception and possible barriers prior to undertaking any treatment^5^. Literature and guidelines cite prognostic estimates that compare various medical treatments to expectant management alone. For example, in couples with unexplained infertility, crude live birth rates of 9-28% could be offered as a prognosis if managed expectantly^6–9^. Many of the couples given this diagnosis will have behavioural, environmental, endogenous and lifestyle factors impacting their fertility that warrant intervention. Thus, many will use concurrent strategies such as fertility awareness methods, weight loss and smoking cessation prior to attempting ART. The synergistic effect of multiple low-risk interventions is not well described in comparison to expectant management.

What are the outcomes if we attempt to treat multiple underlying lifestyle and medical issues from the beginning of the patient journey? Few studies examine birth rates in infertile couples no treated with ART^10^. In this case series, we follow couples seen in a multidisciplinary clinic where they are given lifestyle advice and training in a fertility awareness method after their initial visit. Fertility awareness methods assist couples in identifying the woman’s peak fertile days and assists clinicians in the diagnosis of various disorders and guiding medically assisted reproduction treatments^11,12^. Couples with specific disorders are offered focused interventions including conventional medical treatments, oral ovulation induction and fertility restoring surgery.

This overall approach to infertility has been referred to as ‘restorative reproductive medicine’ (RRM) highlighting its personalised, optimisation focus^13,14^. The outcomes of an RRM-styled clinic have not been described in an Australian context. This study therefore aims to inform clinicians and couples by documenting the outcomes of a restorative approach to infertility.

## Materials and Methods

### Study Population and Setting

We performed a retrospective study on infertile couples attending a hospital-based clinic in Brisbane from January 2005 to December 2010.

We included all couples visiting the clinic and attempting natural conception diagnosed with infertility as defined by the WHO as 12 months of regular, non-contracepted intercourse without conception. Couples that had previously attempted IVF were included. Females who had entered menopause, with bilateral tubal blockage and males affected by azoospermia were excluded.

During the study period the clinic was staffed by General Practitioners (GP) with additional training in fertility awareness and RRM. The clinic receives self-referrals from patients as well as referrals from primary care clinicians or other specialists for treatment of a range of fertility related disorders. The service was not established to offer any ART but does offer medically assisted ovulation induction.

### Fertility Workup and Treatment

After receipt of their referral, couples were contacted by clinic staff for a consultation including preconception health, infertility history, examination and completion of any investigations. At the initial consultation, a plan of treatment was made including inviting couples to undertake fertility awareness charting based on the symptothermal method^15^. This ‘fertility charting’ phase required couples to record basic characteristics of their menstrual cycle including their daily basal body temperature and cervical/vaginal discharge. The use of charting along with focused investigations is an essential aspect of RRM^13^.

After learning fertility tracking over 1-2 months a further investigative phase assesses midluteal progesterone and oestradiol. This often leads to medication use to optimize ovulation, cervical mucus and luteal phase hormone levels. This includes the use of metformin, ovulation induction, and, during those years in the clinic, luteal phase support with progesterone for some couples.

Further personalised medical or surgical treatments by supporting hospital teams were undertaken as clinically indicated according to patient wishes and prognosis. Examples include nutritional, immunological, anti-bacterial and psychological interventions. Tailored treatment plans would proceed in a stepwise fashion until cycle characteristics, hormonal markers and underlying comorbidities had been optimised as much as possible.

### Outcome Measures

The main outcome measure was time to conception resulting in live-birth. Time to conception was calculated as the date from the last menstrual period. Live birth was defined as the birth of a live baby ≥ 22 weeks gestation^16^.

Other outcomes were the rates of clinical pregnancy, miscarriage, ectopic pregnancy and multiple pregnancies. Miscarriage was defined as pregnancy loss before 22 weeks of gestational age^16^.

### Follow-up and Data Collection

Couples were followed for 24 months. Data was collected on patient characteristics, time to conception, date of birth, birth weight, treatments used and infertility diagnostic category (as determined by the treating clinician). Couples declining further care were censored from the analysis at the moment of last contact, and the number of days from initial consultation was calculated.

Data was collected from chart review of clinical records. Data was stored in a locked and deidentified Excel™ (Microsoft Corporation) database by the research team on password-protected computers in the clinic. Follow up of patients was arranged via telephone or email by clinic nurses if not presenting for ongoing care.

### Statistical analysis

We used Kaplan-Meier analysis to assess time-to-event. A survival analysis using the Kaplan-Meier method provides a cumulative live birth rate (CLBR) by censoring for couples discontinuing treatment. This is graphed as a cumulative ongoing pregnancy rate to reflect the delay from pregnancy diagnosis to birth. The distribution of each categorical independent variable was calculated, with the mean or median for continuous variables. Outcomes (resolution of infertility and pregnancy outcome) were further summarized by infertility type. All analyses were conducted in Excel™.

### Ethics

This study received ethical approval by the Mater Hospital Human Research Ethics Committee. Fertility Assessment and Research Clinic Outcomes HREC/MML/44585.

## Results

During the study period, 277 couples presented with the wish to have a child, of which 102 couples were excluded from analysis as they were infertile for less than 12 months. These couples usually had other fertility concerns such as irregular cycles, recent miscarriage after prolonged duration of trying or advanced maternal age. Also, 13 couples were excluded due to incomplete follow up data. Therefore, the analysis is limited to 162 couples (Figure 1).

**Figure 1.**
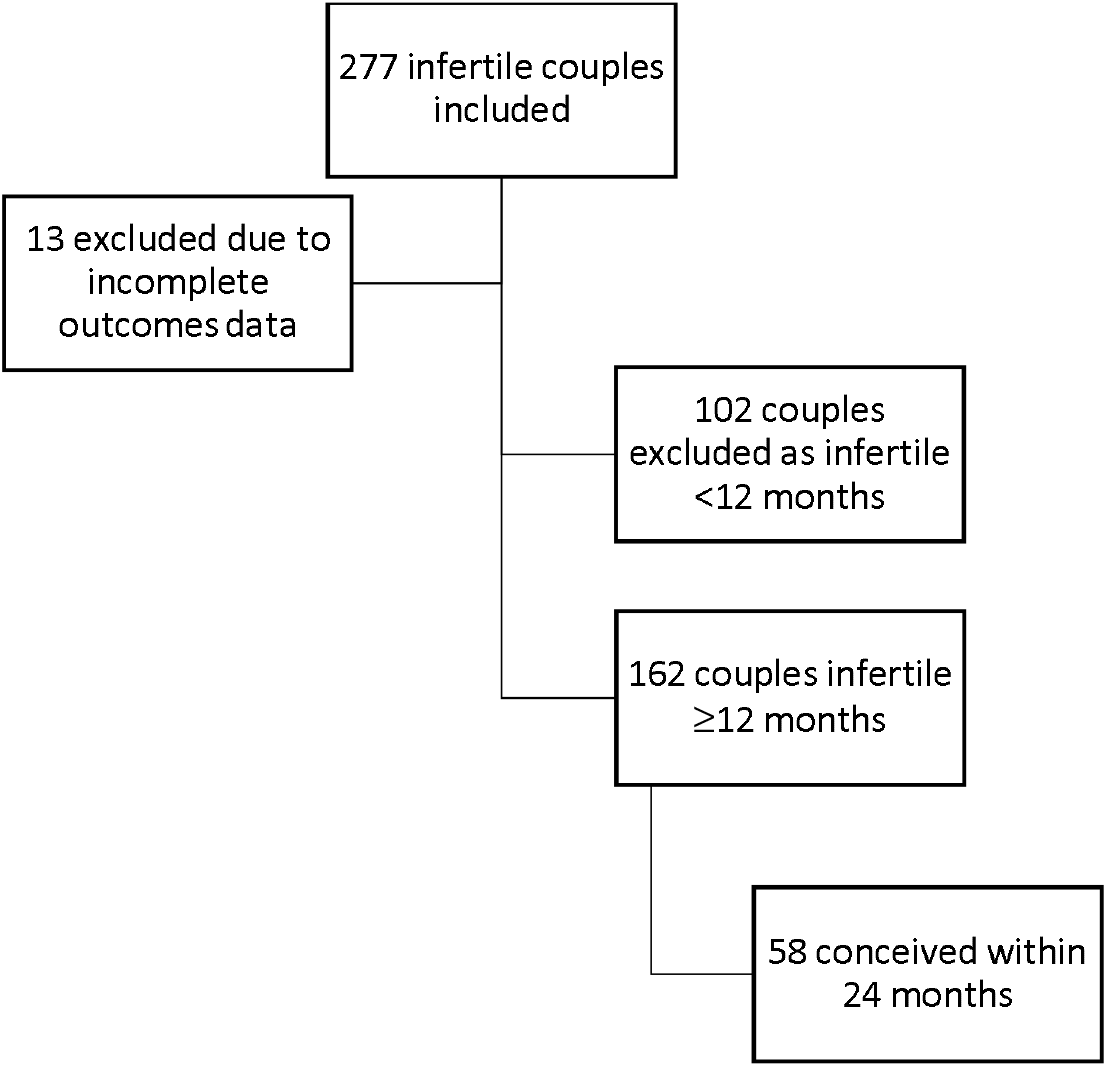
Participant Flow Diagram.

Among these 162 couples, 58 (35.8%) had a conception resulting in live birth within 24 months. The crude clinical pregnancy rate was 44.4% (72/162) while 14 other couples had a pregnancy ending in miscarriage (n=9), ectopic pregnancy (n=4) or termination of pregnancy (n=1; at 21 weeks for MCDA twins with a congenital abnormality).

Using Kaplan-Meier analysis, the CLBR for all couples was estimated to be 10.3% at 3 months, 25.3% at 6 months, 46.0% at 12 months and 57.4% at 24 months (Figure 2). The median time until conception was 5.0 months (range 1-22). The median time follow up for those not conceiving was also 5.0 months (range 1-30) with discontinuation rates of 19% at 3 months, 35% at 6 months and 49% at 12 months. Four couples were known to have proceeded to IUI or IVF whilst still in contact with the treating team. The outcomes of other couples after the study period or after withdrawing from treatment are not known.

**Figure 2.**
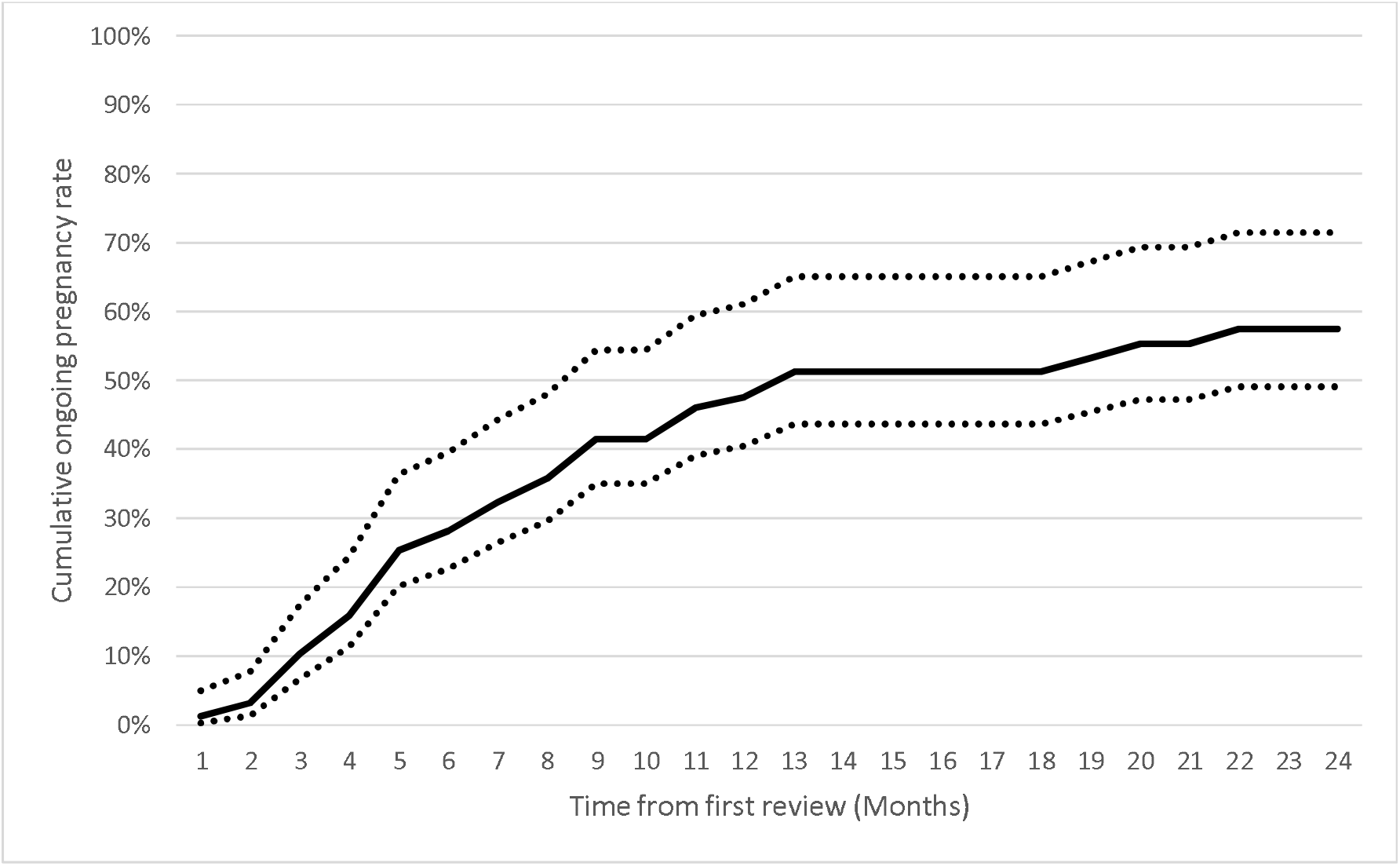
Cumulative ongoing pregnancy rate resulting in live birth for all couples (Kaplan-Meier survival analysis, 95% confidence interval (CI) lines shown. Censored for ceasing treatment with clinic)

All couples being treated were categorised by commonly ascribed causes of infertility (Table 5). The highest pregnancy and birth rates were seen in the ovulatory disorder and unexplained infertility categories, which accounted for 29.6% and 45.7% of couples respectively. The remaining 24.7% of couples had crude birth rates of less than 30%.

**Table 1.** Baseline Characteristics of Participant Couples.

|  | No pregnancy<br>(N=90) | Conceived<br>(N=72) | All<br>(N=162) |
| --- | --- | --- | --- |
| Median female age in years (range) | 33.3 (20-47) | 34.0 (20-44) | 33.7 |
| Median duration of infertility in months (range) | 40 (12-240) | 25.5(12-72) | 33.9 |
| Previous live birth | 31 (34.9%) | 42 (58.1%) | 45.8% |
| Previous IVF/IUI | 12 (13.3%) | 5 (6.9%) | 17(10.5%) |
| Mean body mass index | 26.8 | 26.1 | 26.5 |
| Smoker | 17 (19.3%) | 13 (17.6%) | 30(18.5%) |
| Anovulation/PCOS | 35 (38.6%) | 23 (32.4%) | 58(35.8%) |

**Table 2.** Pregnancy Rates by Female Age Category.

| Female Age Category | ≤30 | >30-35 | >35-40 | >40 |
| --- | --- | --- | --- | --- |
| Total Couples | 48 | 49 | 45 | 20 |
| Live births | 23 | 14 | 17 | 4 |
| Multiples | 1 | 0 | 0 | 0 |
| Miscarriages | 0 | 2 | 7 | 0 |
| Ectopic pregnancies | 3 | 1 | 0 | 0 |
| Terminations of pregnancy | 0 | 1 | 0 | 0 |
| Crude pregnancy proportion | 56.3% | 36.7% | 53.3% | 20.0% |
| Crude live birth proportion | 47.9% | 28.6% | 37.8% | 20.0% |

**Table 3.** Pregnancy Outcomes.

|  |  |
| --- | --- |
| **Total Couples treated** | **162** |
| **Total Confirmed Pregnancies (one per couple)** | 72 |
| **Pregnancy leading to live births** | 58 |
| **Multiple Pregnancies Birthed** | 0 |
| **Miscarriages** | 9 |
| **Ectopic Pregnancies** | 4 |
| **Terminations of pregnancy** | 1<sup>§</sup> |
| **Clinical pregnancy rate** | **44.4%** |
| **Live birth rate** | **35.8%** |
| **Miscarriage rate** | **12.5%** |

<sup>§</sup> Monozygotic-diamniotic twins

Basic birth outcomes were collected. These showed 3 preterm births at a rate of 5.2%. Mean birth weight was 3326 ±623g and median 3355g. The three preterm births were recorded at 24, 31 and 36 weeks. In keeping with this result, only three babies were less than 2500g birth weight. The reasons for preterm birth were not identified. The remaining babies were all normal birth weight or above 4000g at birth.

**Table 4.** Birthweight and Gestation Outcomes.

|  |  |  |  |
| --- | --- | --- | --- |
| **Gestation** | <37 weeks | 3 | 5.2% |
|  | ≥37 weeks | 55 | 94.8% |
| **Birthweight** | <1500g | 1 | 1.7% |
|  | 1500-2500g | 2 | 3.4% |
|  | >2500g-3999 | 47 | 81.0% |
|  | ≥4000g | 8 | 13.8% |

**Table 5.**
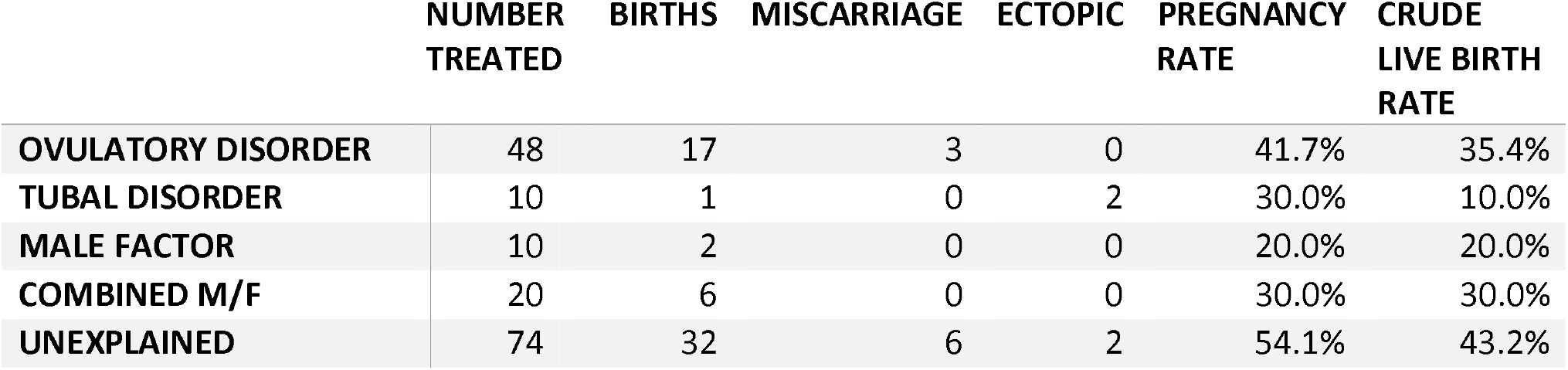
Pregnancy and Crude Live Birth Rates by Infertility Diagnosis.

The categories with the highest crude birth rates were further subjected to a survival analysis. The CLBR for couples with unexplained infertility was 58.7% at 12 months and 71.2% at 24 months. The CLBR for couples with ovulatory disorders was 47.5% at 12 months and 54.0% at 24 months (Figure 3).

**Figure 3.**
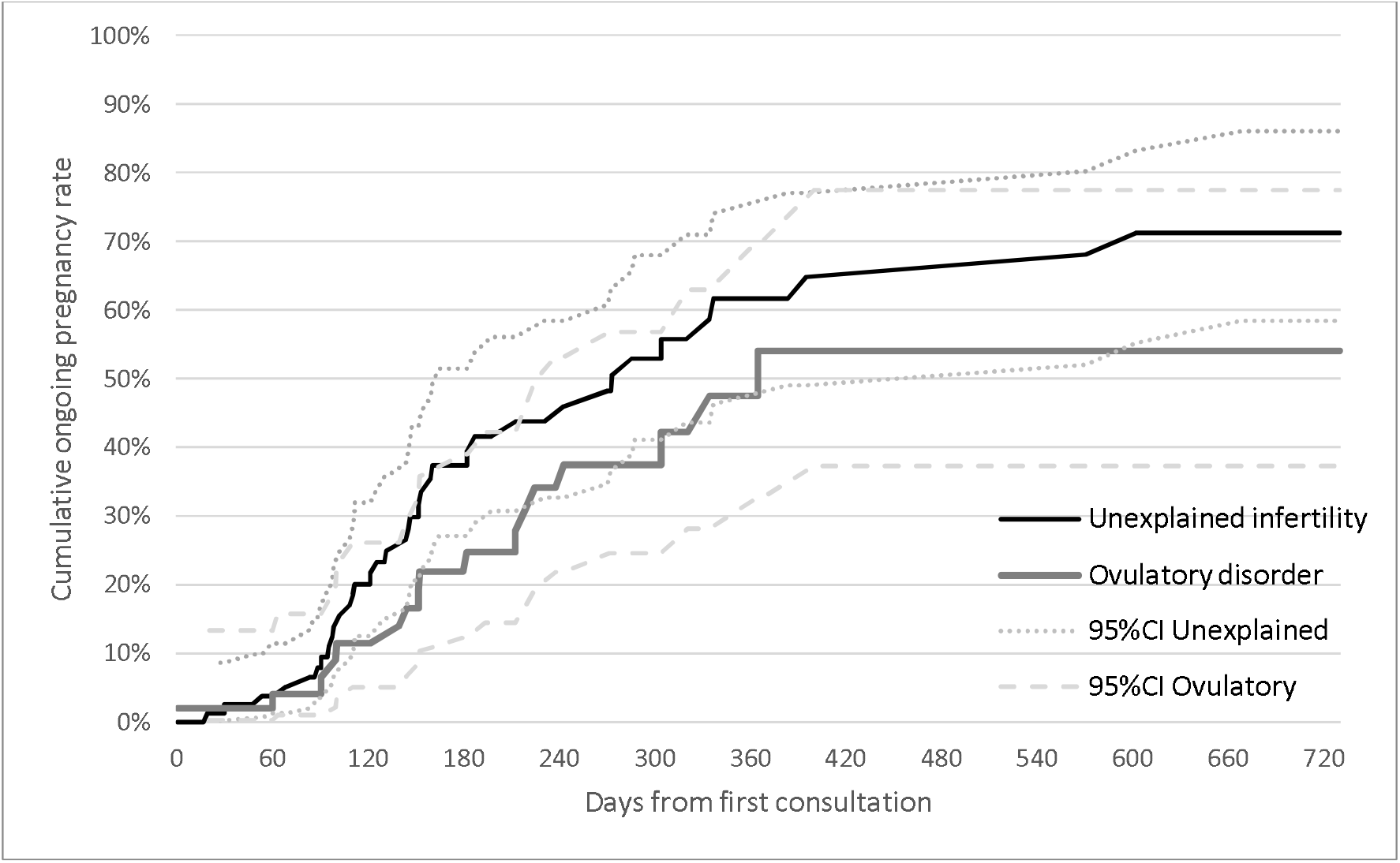
Cumulative ongoing pregnancy rate resulting in live birth for couples with ovulatory disorders or unexplained infertility (Kaplan-Meier survival analysis. Censored for ceasing treatment with clinic)

The treatment approach used in this clinic typically employs multiple interventions individualized to each couple. Specific treatments for every couple were therefore not included in this results analysis due to the complexity of the data, but they included excisional surgery for endometriosis in 8 patients, other fertility related surgeries such as salpingectomy for damaged tube, polypectomy or fibroidectomy in 7 patients, mucus enhancers in 55, luteal phase support with human chorionic gonadotropin or progesterone in 69, ovulation induction in 30, as well as fertility awareness charting, preconception vitamins and folate in all patients.

In addition to recognised causes of subfertility we also recorded multiple other diagnoses that likely impact on fertility such as obesity, diabetes, hypertension, luteal phase defect, clotting disorders, thyroid disorders and factors that impaired optimal timing of intercourse. These conditions were addressed as part of their management wherever possible.

## Conclusions

This study describes the outcomes of a ‘restorative’ treatment approach to infertility in a single Australian secondary centre. The crude live-birth rate of 35.8% and cumulative live-birth rate of 57.4% over 24 months in this case series continues the findings of other studies in this emerging field. The largest prior RRM study examined 1072 couples treated at an Irish general practice. They demonstrated crude and cumulative birth rates of 25.5% and 52.8% respectively over 24 months. This cohort had an older average female age of 35.8 years, a longer average duration of infertility at 5.6 years, with 33% having previously attempted ART^13^. A more recent publication following 108 couples achieved a crude birth rate of 38% and a CLBR of 66% over 24 months. This cohort had an average female age of 35.4 years, a mean infertility duration of 3.2 years, with 22% having attempted intrauterine insemination (IUI) and 9% in-vitro fertilisation (IVF) previously^17^.

Contextualising these outcomes is complicated by the heterogenous patient populations and multifactorial treatment methods. There are several observational reports of heterogenous infertile populations that provide live birth rates over time. One publication collated 20 such studies, following over 14,000 couples, and created a model to predict outcomes based on treatment type. It found that the birth rates of 31% with no treatment, 42% with non-ART treatment and up to 47% if half of the remaining couples proceeded to ART over 3 years^10^. It did not specify duration of infertility, average female age or other important confounders. They did estimate that the non-ART treatment group could expect 7% multiples and the ART 12% multiple births. While no strong causal inference can be made from such observational data, it provides an additional perspective to controlled experimental studies. It provides typical outcomes and types of treatments incorporating the influences of patient choice, economics and ethics.

We describe pregnancy chances cumulatively per started treatment over a defined time frame. Most current literature uses a ‘pregnancy per-cycle’ analysis suited to ART^18^. A time frame of 24 months follow up was chosen as previous publications have shown that cumulative pregnancy rates are only marginally increased after this time frame, irrespective of treatment approach^9^. Cumulative rates by life-table analysis adjust for withdrawals from treatment but may overestimate successes since patients with poor prognosis are perhaps more likely to drop out of treatment^18^.

In fertility prognosis studies, couples are usually categorised by underlying cause and possible treatment modality^10^. The largest two groups in our study were the unexplained infertility and ovulatory disorder groups. The highest birth rates were in the unexplained infertility group with a CLBR of 58.7% at 12 months and 71.2% at 24 months. The crude live birth rate in this group is 39% at 12 months and 43% at 24 months. This figure is higher than published observational studies using expectant management alone for unexplained infertility. For example, one commonly cited paper looking at prognostic models for expectant management in unexplained infertility showed a CLBR of 27% in the 12 months following completion of fertility investigations^19^. Another study of 3021 ‘good prognosis’ infertile couples found a CLBR of 29.5% over 12 months^8^.

Comparison of our case series with expectant management has multiple limitations as most fertility studies consider expectant management as ‘no treatment’. By contrast, couples in this restorative treatment series had ongoing fertility charting guidance, lifestyle advice and may have been referred to other specialists for medical optimisation or infertility surgeries.

Our couples with a poor prognosis such as tubal and male factor infertility groups had very low birth rates consistent with other non-ART studies. For example, one study found a CLBR of 2.4-6.6% for patients with a tubal factor, severe male factor or longstanding unexplained infertility whilst waiting for IVF^20^.

A common question by those not familiar with the RRM approach to infertility is how long does treatment take? In our series the median time to pregnancy was 5.0 months. Comparing time-to-conception with other heterogenous cohorts is difficult, particularly when including couples managed with episodic ART in the era of frozen transfer cycles. For reference, one local cohort study of 1386 couples at a single infertility centre in New Zealand showed an average of 2.4 years to achieve pregnancy for those requiring IVF, 1.7 years for non-IVF treatment and 1.4 years if successful with expectant management^21^. Interestingly, this cohort was followed for 13 years. During that time 35% of couples ultimately required ‘treatment’ to achieve pregnancy, but 27% spontaneously conceived within 13 years of follow up. Another study focusing on unexplained infertility described an overall time to pregnancy of 8.1 months in a good prognosis group versus 14.0 months in a poor prognosis group after stratification and treatment with expectant management or ART^9^.

We have reported very basic pregnancy outcomes including twin deliveries (0.0%), miscarriage (12.5%), ectopic pregnancies (5.6%) and low preterm birth rates of 5.2%. The overall numbers in each of these categories are too small to warrant any further analysis.

Our discontinuation rates are comparable to the 50% discontinuation rate in the Irish RRM study. It is higher than the 22.9% rate in a Dutch publicly funded fertility clinic cohort^22^. In other studies, IVF treatment discontinuation rates are 17-70% when not publicly funded^22^. Prior studies show older couples and those with secondary infertility are most likely to discontinue. Other reasons include a dislike of the care offered, emotional, financial or relational issues^23^. We did not follow long-term outcomes of patients who discontinued care.

In summary, this case series defines outcomes from a distinctive fertility treatment approach using multidisciplinary care focusing on restoring fertility health. Our study’s main limitations are its small, observational, statistically unadjusted and retrospective nature. It demonstrates similar outcomes to prior RRM studies, thus supporting its generalisability. As an observational study with broad inclusion criteria and multiple common interventions, this data offers insight into the possible outcomes for couples seeking comprehensive non-ART care. It also highlights the need for ongoing data collection of all fertility therapies^24^ in an evolving era of precision and personalized medicine.

## Data Availability

This data is available through contacting the main author or the Fertility and Research clinic at the Mater Hospital, Brisbane.

